# Schizandrin A suppressed pancreatic cancer proliferation and cancer associated fibroblast activation via inhibiting the COX-2/ALOX5 shunting

**DOI:** 10.1101/2024.10.09.24315180

**Authors:** Zhirui Zeng, Shan Lei, Jingya Wang, Dahuan Li, Yushi Yang, Qianting Tian, Xiaojiang Hao, Tengxiang Chen

## Abstract

**Background:** Schizandrin A is major components extracted from *Schisandra chinenzis-Turcz. Baill* and *Schisandra sphenanthear Rend. etWils*. Schizandrin A exhibits remarkable hepatoprotective, antiviral and anti-inflammatory effects. However, the anti-tumor effects and its molecular mechanism were still known limited.

**Methods:** The affinity between Schizandrin A and COX-2/ALOX5 protein was analyzed using network pharmacology, computer molecular docking, and surface plasmon resonance experiments. Bioinformatic analysis and review of clinical characteristics were conducted to assess the necessity of simultaneous blocking of COX-2 and ALOX5 in pancreatic cancer (PC). LC/MS metabolomics and RNA-sequencing were utilized to investigate the effects of schizandrin A on the activation and expression of COX-2/ALOX5 in PC cells. Biological function experiments were conducted to investigate the inhibitory effects of Schizandrin A on PC cell proliferation and cancer-associated fibroblast activation *in vitro* and *in vivo*.

**Results:** Schizandrin A demonstrated a high affinity for binding directly with COX-2 and ALOX5, with kinetic association constants of 14.8 μM and 21.8 μM, respectively. PC exhibited a significant COX-2/ALOX5 signature, while PC cases with a high COX-2/ALOX5 signature showed lower overall survival and disease-free survival rates. Treatment of PC cells with schizandrin A resulted in decreased COX-2/ALOX5 activity and expression, leading to inhibition of leukotriene and prostaglandin production, as well as suppression of the downstream pathway NF-kappaB signaling. Schizandrin A demonstrated significant inhibitory effects on the proliferation and sphericity of PC cells *in vitro*, as well as on cell proliferation *in vivo*, while exhibiting low toxicity to normal tissues. Treatment of conditioned medium from PC cells with schizandrin A resulted in reduced induction of normal fibroblasts into cancer-associated fibroblasts. Furthermore, mutations in the binding sites of ALXO5 (Arg246) and COX-2 proteins (Ile124 and Ser126) resulted in a significant decrease in affinity to Schizandrin A, and blocking the inhibitory effects of schizandrin A.

**Conclusions:** Taken together, schizandrin A directly bound with COX-2 and ALOX5, reduced their activation and leukotrienes and prostaglandins production, thus exhibiting distinguished effects on suppressing PC proliferation and inhibiting the ability of PC cell to induce normal fibroblasts to transform into tumor-associated fibroblasts. Therefore, schizandrin A represents a potentially novel therapeutic approach for PC.

## 1. Introduction

Pancreatic cancer (PC) is a highly aggressive malignancy of the digestive tract characterized by delayed diagnosis, frequent metastasis, poor prognosis, and resistance to chemotherapy [1]. The five-year survival rate for patients with PC is less than 5%. Despite advancements in surgical techniques and chemotherapy regimens, the overall prognosis for PC patients remains bleak [2,3]. The microenvironment of PC is characterized by an abundance of growth factors and cytokines that promote tumor growth and induce immunosuppression. Research has demonstrated that up-regulation of cytokine pathways plays a crucial role in regulating the progression of PC and evading immune responses [4,5]. Consequently, targeting cytokine production may offer potential therapeutic advantages in the treatment of PC. Prostaglandins (PGs) and leukotrienes (LTs) are important cytokines involved in promoting tumor progression, which can be produced by tumor cells or stromal cells through the precursor arachidonic acid (AA) [6]. Cyclooxygenase, lipoxygenase, and cytochrome P450 are the rate-limiting enzymes involved in AA metabolism [7]. Cyclooxyganese-2 (COX-2), a key member of the cyclooxygenase family, catalyzes the conversion of AA into PGs and TXA2 [8]. In tumor tissues, COX-2 continuously induces the production of cytokines and promotes the activation of NF-kappaB signaling, thus leading to tumor progression [9,10]. To date, several COX-2 inhibitors, such as celecoxib and rofecoxib, have been identified. These COX-2 selective inhibitors suppress the activation of various pathways by reducing the production of cytokines in tumor tissues, thereby inhibiting tumor progression [11]. In addition, COX-2 selective inhibitors have synergistic effects with a variety of chemotherapeutic drugs, including gemcitabine, cisplatin, and 5-FU [12]. However, some clinical trials have shown that the inhibitory effect of COX-2 selective inhibitors on suppressing cytokine production in tumor tissues is gradually weakened after prolonged use, requiring a gradual increase in dose. However, severe cardiac, gastrointestinal, and renal toxicity in patients with cancer limits the application of high-dose COX-2 inhibitors [13]. AA is a common mediator of COX-and LOX-induced cytokine production; a signaling shunt from COX-2 to LOX is observed in COX2 inhibitor-resistant cancer. COX-2 deficiency promotes feedback for the activation and expression of ALOX-5, which still induces cytokine production by utilizing AA and rescues the inhibitory effect of the COX-2 inhibitor [14,15]. Therefore, drugs targeting COX-2/ALOX5 shunting may contribute to cancer therapy.

Natural products refer to the components or metabolites of animals, plant extracts, and insects. Various active components of natural products have structural diversity and biological activity, which play a pharmacological role by targeting multiple targets and can be used as a molecular library for multi-target drug screening [16]. Schizandrin A is a major component extracted from *Schisandra chinenzis-Turcz. Baill* and *Schisandra sphenanthear end. EtWils* [17]. Previous studies have indicated that Schizandrin A exhibits remarkable hepatoprotective, antiviral, and anti-inflammatory effects [18]. In our previous study, we found that schizandrin A exhibits hepatoprotective effects and predicted to inhibit AA metabolism [19]. However, the precise target of Schizandrin A remains to be elucidated.

In the current study, we demonstrated that schizandrin A directly bound with COX-2 and ALOX5, reduced their activation and the production of leukotrienes and prostaglandins, thus exhibiting distinguished effects on suppressing PC cell proliferation and inhibiting the ability of PC cell to induce normal fibroblasts to transform into tumor-associated fibroblasts. Therefore, schizandrin A represents a potentially novel therapeutic approach for PC.

## 2. Materials and methods

### 2.1. Cell lines, cell culture, and chemicals

Normal human pancreatic epithelial cells (HPEC), normal fibroblast (NF), 293T and pancreatic cancer cells (PA-TU-8988T, Panc0327, SW1990, CFPAC-1, Mia-PaCa2, Panc1005, BXPC-3 and HPAC) were all obtained from Procell (Wuhan, China). All PC cells were cultured in Dulbecco’s modified Eagle’s medium (DMEM; Gibco, USA) containing 10% fetal bovine serum (FBS; Bioind, Israel). HPEC, NFs and 293T were cultured in Roswell Park Memorial Institute-1640 (RPMI-1640; Gibco, USA) medium containing 10% FBS. All the cells were cultured at 37 °C with 5% CO2. Schizandrin A (Cat. No. HY-N0693) was obtained from MedChemExpress (Wuhan, China). Schizandrin A were dissolved in DMSO, adjusted to a concentration of 10 mM, and stored at -20 °C prior to use.

### 2.2. Plasmids and transfection

The full-length sequences coding for ALOX5 and COX-2 were cloned into the pReceiver-Lv105 expression vector to construct wild-type (WT) ALOX5 and COX-2 overexpression plasmids. The mutant (MUT) COX-2 plasmids were constructed using alanine (Ala) to replace 124 isoleucine (124 Ile) and 126 serine (126 Ser), respectively, while the ALOX5 MUT plasmid was constructed using alanine (Ala) to replace 246 arginine (246 Arg). All plasmids were constructed by Beijing Viewsolid Biotech Co., Ltd. Lipofectamine 2000 (Thermo Fisher Scientific, USA) was used to transfect plasmids into cells according to the manufacturer’s instructions.

### 2.3. Network pharmacology and computer molecular docking

First, the potential targets of schizandrin A were predicted using online tools Swissprediction (http://www.swisstargetprediction.ch/) and TargetNet (http://targetnet.scbdd.com/calcnet/index/). The structure of schizandrin A was obtained from PubMed Compound (https://www.ncbi.nlm.nih.gov/pccompound). The crystal structures of PTGS2 (ID: 5f1a), ALOX5 (ID: 6n2w), PTGS1 (ID: 6y3c), CHRM1 (ID: 6wjc), OPRK1 (ID: 6vi4), CYP3A4 (ID: 4d7d), PDE10A (ID: 4bbx), HSD11B1 (ID: 2bel), CDK2 (ID: 1b38), PDE4D (ID: 7cbq), CYP2C19 (ID: 4gqs), DYRK1A (ID: 6a1f), KCNH2 (ID: 6syg), TYK2 (ID: 7k7o), and PDE7A (ID: 4y2b) were obtained from the Protein Data Bank (https://www.rcsb.org/). Then, schizandrin A and target structures were imported to the SYBYL-X (version 2.1) to perform flexible docking according to the standard procedure. The binding sites of the targets and Schizandrin A were visualized using Pymol software (version 1.1).

### 2.4. Surface plasmon resonance (SPR)

Purified ALOX5 and COX-2 protein were desalinated using the AKTA protein purification system (GE Healthcare, USA). Then, the proteins were dissolved in buffer solution (200 mM HEPES, 2 mM NaCl and 0.5% DMSO) and combined with sodium acetate solution (pH 4.0) and CM5 chip in Biacore T200 (GE Healthcare, USA). Following conjugation with hMOF in the CM5 chip, ethanolamine solution was used to block the uncoupled proteins. The chip was then added to the buffer solution for 10 h. Schizandrin A was dissolved in the buffer solution and set as the mobile phase. A multi-cycle model was used to detect the binding constants between the proteins and Schizandrin A. The flow rate of the mobile phase was set as 30 μL/s, the binding time was set to 240 s, and the dissociation time was set to 300 s.

### 2.5. Tissue ethical statement and immunohistochemistry (IHC)

The PC tissues and their corresponding adjacent tissues utilized in this study were procured from the Department of Hepatobiliary Surgery at Guizhou Medical University in accordance with the principles outlined in the Declaration of Helsinki. Informed consent was obtained from all patients who contributed samples, and the collection and utilization of these tissue specimens were sanctioned by the Human Research Ethics Review Committee at Guizhou Medical University. For IHC, after permeabilizing, dehydration, and antigen retrieval, 0.3% H2O2 and 5% BSA were used to block the sections. Then, the sections were stained with antibodies against COX-2 (1:100, Cat no. 27308-1-AP, Proteintech, Wuhan, China), ALOX5 (1:100, Cat no. A2877, Abconal, Wuhan, China), Ki67 (1:200, Cat no. 27309-1-AP, Proteintech, Wuhan, China), PCNA (1:200, Cat no. 10205-2-AP, Proteintech, Wuhan, China), α-SMA (1:100, Cat no. 55135-1-AP, Proteintech, Wuhan, China), and FAP (1:160, Cat no. #52818, CST, USA) for 12 h at 4 °C. After washing with PBS three times, horseradish peroxidase-conjugated secondary antibodies were used to stain sections, and 3,3-diaminobenzidine was stained to detect the immunologic signals. After using hematoxylin to stain the nucleus, the tumor sections were imaged using a light orthophoto microscope (magnification ×200, Olympus, Japan).

### 2.6. Liquid chromatography/mass spectroscopy (LC/MS) metabonomics

All samples were thawed at 4°C and transferred into 2 mL centrifuge tubes. A total of 100 μl 2-chlorobenzalanine (4 ppm) methanol solution was added, and samples were vortexed for 5 min. After centrifuging at 4 °C for 10 min at 12000 rpm, the samples were filtered through a 0.22 µm membrane to obtain samples for LC-MS. LC/MS was performed using a Thermo Vanquish (ThermoFisher, USA) equipped with an ACQUITY UPLC® HSS T3 (150 × 2.1 mm, 1.8 µm, Waters) column maintained at 40 °C. The temperature of the autosampler was maintained at 8 °C. The ESI-MSn experiments were performed on a Thermo Q Exactive Focus mass spectrometer (ThermoFisher, USA) with a spray voltage of 3.8 kV and -2.5 kV in positive and negative modes, respectively. Dynamic exclusion was performed to remove unnecessary information from MS/MS spectra. The differential metabolites were analyzed using MetaboAnalyst 5.0, while metabolites with |FC ≥ 2| and P-value < 0.05, were differentially expressed metabolites.

### 2.7. RNA sequencing

Total RNA was extracted from the samples using TRIzol reagent (ThermoFisher, USA) following the manufacturer’s instructions. RNA quantity and purity were analyzed using a Bioanalyzer 2100 and RNA 6000 Nano LabChip Kit (Agilent, CA, USA), and high-quality RNA samples with an RIN number of > 7.0, were used to construct the sequencing library. Following purification, the mRNA was fragmented into short fragments and reverse-transcribed to generate cDNA using SuperScript™ II Reverse Transcriptase (Invitrogen, USA). The average insert size of the final cDNA library was 300 ± 50 base pairs (bp). Finally, the cDNA was sequenced on an Illumina Novaseq 6000 (LC-Bio Technology Co., Ltd., Hangzhou, China) following the manufacturer’s protocol. After obtaining high-quality clean reads and batch normalization, the count data were analyzed using the EdgeR package. Genes with LogFC ≥ 1 and P-value < 0.05 were set as differentially expressed genes.

### 2.8. ELISA experiments

The levels of prostaglandin A2 (PGA2) and leukotriene F4 in PC cells were detected by ELISA experiments. PC cells after indicated treatment were lysated by RIPA buff, and the levels of PGA2 and leukotriene F4 in each group cells were determined using Prostaglandin A2 ELISA Kit (Biorbyt, England) and LT-F4 GENLISA™ ELISA (KRISHGEN, India) according to the manufacturer’s specification, respectively. The levels of PGA2 and leukotriene F4 were normalized to protein concentration.

### 2.9. Quantitative real-time PCR (qRT-PCR)

Total RNA was isolated from PC cell lines utilizing TRIzol® reagent (Invitrogen, USA), followed by cDNA synthesis using the 1st Strand cDNA Synthesis Kit (Invitrogen, USA) according to the manufacturer’s protocol. qRT-PCR was performed using SYBR green (Invitrogen, USA)under the following conditions: initial denaturation at 95°C for 5 min, followed by 40 cycles of denaturation at 95°C for 30 s, annealing at 60°C for 30 s, and elongation at 72°C for 30 s. β-actin served as the internal control for normalization. Primers used in the current study were showed as followed: CSNK2B forward primer 5’-TGAGCAGGTCCCTCACTACC-3’ and CSNK2B reverse primer 5’-GTAGCGGGCGTGGATCAAT-3’; TLR4 forward primer 5’-AGACCTGTCCCTGAACCCTAT-3’ and TLR4 reverse primer 5’-CGATGGACTTCTAAACCAGCCA-3’; TNFAIP3 forward primer 5’-TCCTCAGGCTTTGTATTTGAGC-3’ and TNFAIP3 reverse primer 5’-TGTGTATCGGTGCATGGTTTTA-3’; TRAF5 forward primer 5’-CCACTCGGTGCTTCACAAC-3’ and TRAF5 reverse primer 5’-GTACCGGCCCAGAATAACCT-3’; ZAP70 forward primer 5’-CGAGCGTGTATGAGAGCCC-3’ and ZAP70 reverse primer 5’-ATGAGGAGGTTATCGCGCTTC-3’; β-actin forward primer 5’-CATGTACGTTGCTATCCAGGC-3’ and β-actin reverse primer 5’-CATGTACGTTGCTATCCAGGC-3’. The expression of target genes were normalized to β-actin.

### 2.10. Western blotting

All cells were homogenized in high-efficiency RIPA lysis reagent (Boster Biological Technology Co. Ltd., Wuhan, China) containing 1% proteasome inhibitor PMSF (Boster Biological Technology Co. Ltd., Wuhan, China). Total protein in cells was collected via a high-speed centrifugation method, and the concentration was determined using the bicinchoninic acid method (Beyotime, Jiangsu, China). Followed by adding the loading buffer, the protein samples (40 μg/sample) were electrophoresed and then transferred to the PVDF membranes (Thermo Fisher, USA). After blocking with the skimed milk powder, the primary antibodies including anti-COX-2 (dilution 1:500; Cat no. 27308-1-AP, Proteintech, Wuhan, China), anti-ALOX5 (Dilution 1:500; Cat no. 10021-1-Ig, Proteintech, Wuhan, China) and anti-β-actin (ACTB: dilution 1:5000; Cat no. AC026, Abconal, Wuhan, China) were added to the membranes at 4 °C overnight. After washing three times with TBST and incubating with secondary antibodies for 2 h at room temperature, an enhanced ECL reagent (Boster, Wuhan, China) was used to visualize the bands. The relative expression of COX-2, and ALOX5 was normalized to ACTB.

### 2.11. Cell count kit-8

Cells were seeded onto a 96-well plate at a density of 3000 cells/well and cultured at 37 °C for 48 h. Then, the medium was removed and 100 μL of fresh DMEM medium containing 10 μL of CCK-8 reagent (Boster, Wuhan, China) was added. After culturing at 37 °C for 2 h, the absorbance of each well was detected in a multiscan spectrum (Bio-Rad, USA) at 450 nm.

### 2.12. 5-Ethynyl-2’-deoxyuridine (EdU)

A total of 2 × 10^5^ cells were seeded on confocal dishes and cultured at 37°C until cell adherence. Then, the cells were incubated with the EdU reagent (Beyotime, Suzhou, USA) and treated with Schizandrin A or DMSO for 4 h. Then, the cells were fixed with 4% paraformaldehyde and permeabilized with 0.3% Triton-X (Servicebio, Wuhan, China). After washing with PBS three times, a click reagent was added to enhance the fluorescence of the EdU reagent. After staining with DAPI to visualize the cell nuclei, the cells were imaged using a fluorescence microscope (ZEISS, Germany).

### 2.13. Colony formation assay

PC cells were seeded in six-well plates at a concentration of 1 × 10^3^ cells per well and subsequently cultured with indicated conditions. Following incubation period for 14 days, the cell colonies were fixed using PFA and stained with 0.5% crystal violet (Boster, Wuhan, China). Subsequently, the colony formation plates were imaged and quantified.

### 2.14. Sphere-forming experiment

Cells were collected and resuspended in DMEM containing 10% FBS. Then, the cells were seeded onto a 96-well ultra-low adsorption microporous plate (Corning, USA) at a density of 1000 cells/well. After spheroid formation, the change in the diameter of the spheroids was detected every two days.

### 2.15. In vivo experiment

The animal experiments were approved by the Animal Ethics Committee of the Guizhou Medical University. Female BALB/c nude mice (4–6 weeks old) were obtained from the Experimental Animal Center of Guizhou Medical University. The mice were fed in the specified pathogen-free environment with a 12 h light/12 h dark cycle. SW1990 cells were resuspended at a density of 1 × 10^7^ cells/mL, and 200 μl PC cell suspension which mixed with or without 1 ×10^5^ NFs was injected subcutaneously into the right flank of mice. On day 9, tumor size was determined, and mice with a tumor size of 40–60 mm^3^ were selected for further experiments. Mice were intraperitoneally injected with DMSO or Schizandrin A (40 mg/kg) every three day, while tumor size was calculated every three days using the following equation: (length × width^2^)/2. Once the diameter of the tumor reached 12 mm, mice were sacrificed, and the kidneys, liver, heart, intestine, and tumor tissues in mice were collected for further experiments.

### 2.16. Immunofluorescence

Following the specified treatment, PC cells were subjected to a series of steps including washing with PBS, fixation with 4% paraformaldehyde for 20 min, and permeabilization using 0.1% Triton X-100 for 10 min. Subsequent procedures involved additional washing with PBS, incubation with the suitable primary antibody including α-SMA (1:50, Cat no. 55135-1-AP, Proteintech, Wuhan, China), and FAP (1:100, Cat no. #66562, CST, USA) in a humidified chamber at 4°C, followed by overnight incubation, washing, and incubation with the appropriate secondary antibody for 2 hours at room temperature. Following thorough washing, the cells were counterstained with DAPI for 8 minutes. After washing by PBS, the immune signal was visualized.

### 2.17. Statistical analysis

The results of the current study were analyzed using SPSS (version 19.0). Differences between two groups were analyzed using Student’s t-test, while differences between multiple groups were analyzed using one-way ANOVA combined with Bonferroni post-test. Statistical significance was set at P < 0.05.

## 3. Results

### 3.1. Schizandrin A had a high affinity to COX-2 and ALOX5 protein

In order to accurately analyze the role of schisandrin A, network pharmacology was performed using Swissprediction and TargetNet online tool to explore the potential targets of schizandrin A (Fig. 1A). A total of 15 potential protein targets of schizandrin A were identified (Fig. 1B-C). We then used schizandrin A for computer molecular docking. Particularly, the docking scores between schizandrin A and COX-2/ALOX5 were the highest among these targets (Fig. 1D). In detail, the methoxy group of Schizandrin A acted as a hydrogen bond acceptor to form two hydrogen bonds with Ile124 and Ser126 of the COX-2 B chain at distances of 1.7 Å and 2.1 Å, respectively (Fig. 1E). As for the binding between Schizandrin A and ALOX5, the methoxy group of Schizandrin A acted as a hydrogen bond acceptor to form a hydrogen bond with Arg246 of the ALOX5 B chain at a distance of 2.0 Å. Moreover, the benzene ring of Schizandrin A and Arg370 of ALOX5 formed a cation-π bond (Fig. 1F). Similarly, SPR results exhibited that the kinetic association constant (KD) of Schizandrin A for COX-2 and ALOX5 was 14.8 and 21.8 μM (Fig. 1G). Taken together, these evidences indicated that schizandrin A had a high affinity to COX-2 and ALOX5.

**Figure 1.**
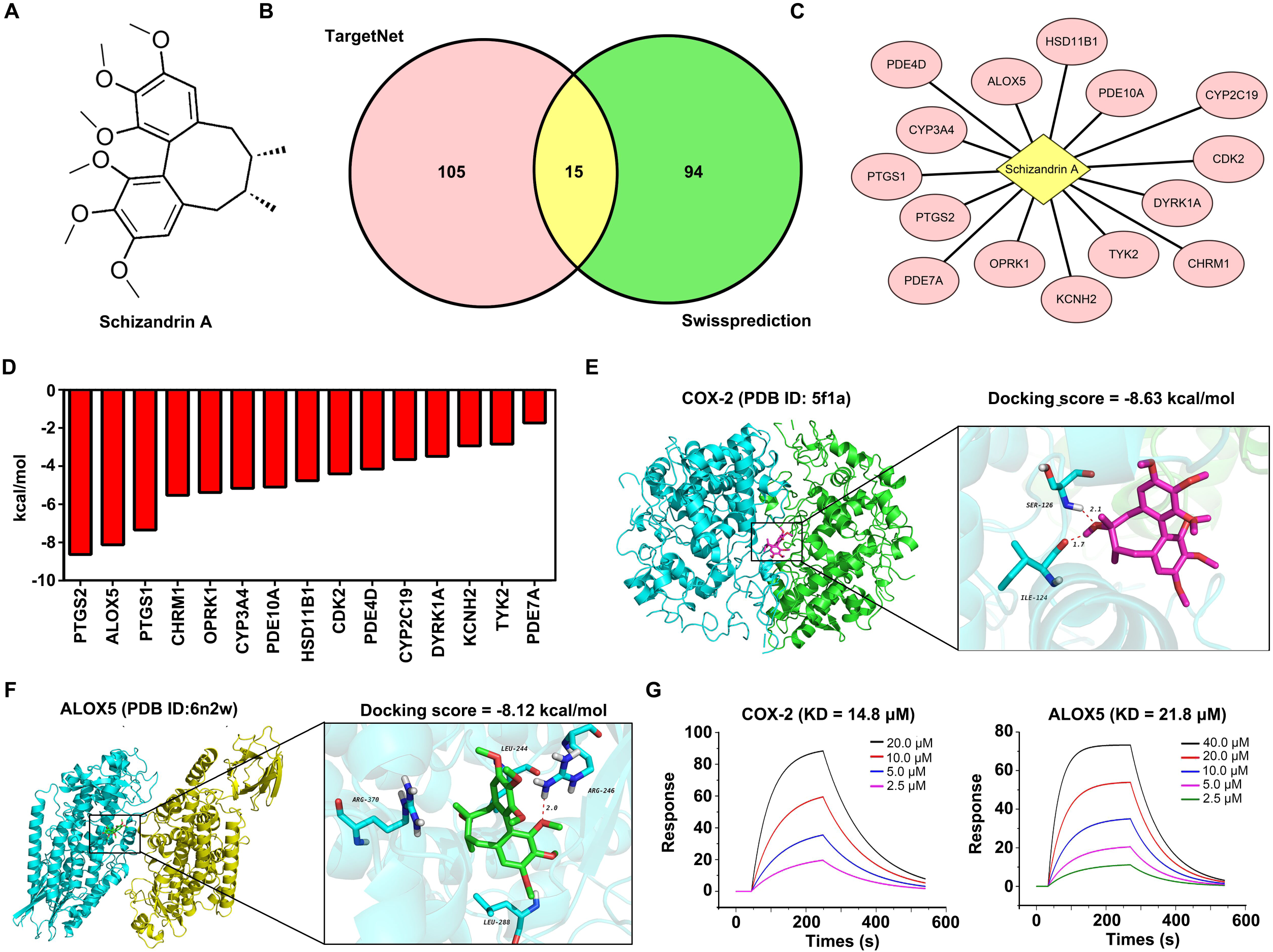
Schizandrin A had a high affinity to COX-2 and ALOX5 protein. (A) Chemical construction of schizandrin A. (B-C) Online tools TargetNet and Swissprediction were used to predict potential targets of Schisandrin A. (D) The docking score of Schisandrin A and potential targets. (E) 3D mode patterns demonstrating the binding sites and binding model of Schizandrin A and COX-2 protein. (F) 3D mode patterns demonstrating the binding sites and binding model of Schizandrin A and ALOX5 protein. (G) Surface plasmon resonance exhibiting the association constant of Schizandrin A, COX-2, and ALOX5.

### 3.2. PC was necessary to concurrently block COX-2 and ALOX5

As schizandrin A had a high affinity to COX-2 and ALOX5, we then analyzed which cancer exhibited up-regulated these targets. COX-2/ALOX5 combined signature was analyzed in pan-cancer in TCGA database, and high level COX-2/ALOX5 combined signature was exhibited in esophageal carcinoma (ESCA), acute myeloid leukemia (LAML) and pancreatic cancer (PAAD) tissues in comparison to their corresponding non-tumor tissues (Fig. 2A). In the PAAD cohort of TCGA database, patients with high COX-2/ALOX5 combined signature exhibited lower overall survival days (Fig. 2B) and disease free survival days (Fig. 2C). These evidences indicated that schizandrin A may be suitable for PC therapy.

**Figure 2.**
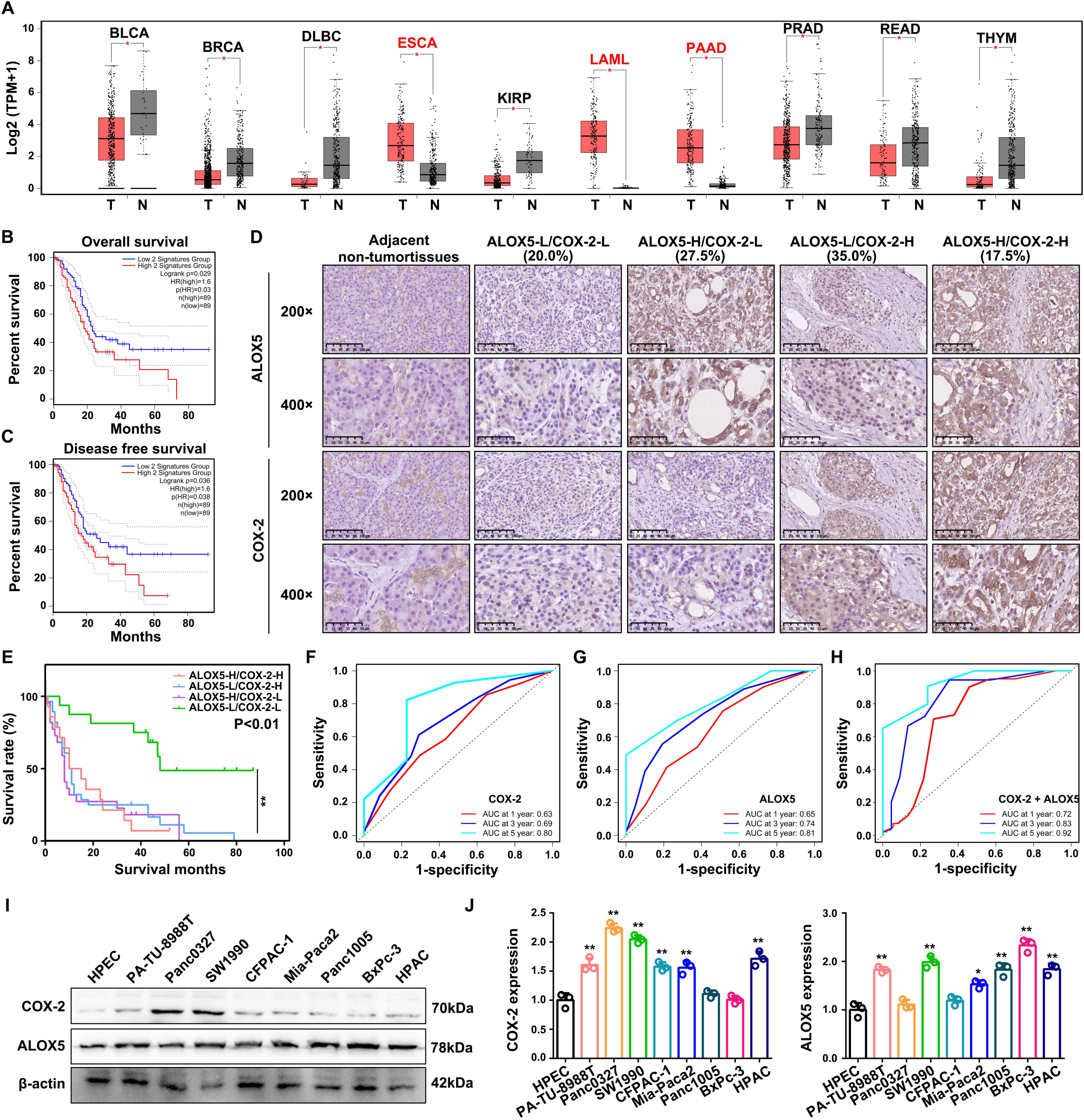
PC was necessary to concurrently block COX-2 and ALOX5. (A) COX-2/ALOX5 signature in pan-cancer tissues in TCGA cohort was calculated, as well as corresponding non-tumor tissues. (B) High COX-2/ALOX5 signature was predicted lower overall survival rate in PC patients of TCGA cohort. (C) High COX-2/ALOX5 signature was predicted lower disease free survival rate in PC patients of TCGA cohort. (D) IHC assays were used to detect the protein expression of COX-2 and ALOX5 in 80 pair PC tissues and adjacent tissues. (E) Subgroup analysis revealed no significant difference between patients with up-regulation of both ALOX5 and COX-2 compared to those with up-regulation of only ALOX5 or COX-2. (F) Diagnostic value of COX-2 for predicting PC patient survival. (G) Diagnostic value of ALOX5 for predicting PC patient survival. (H) Diagnostic value of combination of COX-2 and ALOX5 for predicting PC patient survival. (I-J) The expression of COX-2 and ALOX5 in PC cells and HPEC cells. *, P<0.05; **, P<0.01.

Moreover, we used IHC to detect the expression of COX-2 and ALOX5 in 80 pair PC tissues and corresponding adjacent tissues. Based on the findings, it was determined that 27.5% of PC patients exhibited higher levels of ALOX5, while 35% exhibited higher levels of COX-2 (Fig. 2D). Additionally, 17.5% of PC patients showed up-regulation of both ALOX5 and COX-2 (Fig. 2D). Subgroup analysis revealed no significant difference between patients with up-regulation of both ALOX5 and COX-2 compared to those with up-regulation of only ALOX5 or COX-2 (Fig. 2E). However, patients with low expression of both ALOX5 and COX-2 had significantly longer survival times compared to PC patients in other groups (Fig. 2E). The prognostic significance of COX-2 protein in predicting survival at 1, 3, and 5 years was found to be 0.63, 0.69, and 0.80, respectively (Fig. 2F). Similarly, the prognostic values of ALOX5 protein for predicting survival at 1, 3, and 5 years were determined to be 0.65, 0.74, and 0.81 (Fig. 2G). Furthermore, the combined prognostic values of COX-2 and ALOX5 proteins for predicting survival at 1, 3, and 5 years were calculated to be 0.72, 0.83, and 0.92, respectively (Fig. 2H). Moreover, the expression of COX-2 and ALOX5 proteins was identified in HPEC and PC cell lines, with a notable up-regulation of ALOX5 or/and COX-2 proteins observed in most PC cell lines compared to HPEC cells (Fig. 2I-J). These findings underscore the significance of simultaneous inhibition of COX-2 and ALOX5 in the treatment of PC.

### 3.3. Schizandrin A inhibited the biological functions of ALOX5/COX-2 in PC cells

Based on the aforementioned evidences, we conducted an analysis to investigate the potential inhibitory effects of schizandrin A on the biological functions of ALOX5/COX-2 in PC cells. Subsequently, LC/MS metabonomics was utilized to assess metabolic changes in PC cells following treatment with schizandrin A. The results revealed distinct metabolic patterns in schizandrin A treated PC cells compared to DMSO (Fig. 3A). Specifically, PC cells treated with schizandrin A exhibited 70 down-regulated metabolites and 169 up-regulated metabolites compared to those treated with DMSO (Fig. 3B). Enrichment analysis revealed that out of the 70 down-regulated metabolites, 8 metabolites, including 8(S)-HETE, leukotriene F4, prostaglandin A2 (PGA2), prostaglandin F2a, leukotriene E4, 20-Carboxy-leukotriene B4, 5-KETE, and N-Acetyl-leukotriene E4, were significantly enriched in arachidonic acid (AA) metabolism (Fig. 3C). Among these metabolites, prostaglandin A2 and prostaglandin F2a were found to be catalyzed by COX-2, while the remaining metabolites were catalyzed by ALOX5. Consistent with the findings of LC/MS metabonomics, ELISA experiments revealed that treatment of PC cells with schizandrin A resulted in decreased levels of PGA2 and leukotriene F4. Additionally, a reduction in the protein levels of both COX-2 and ALOX5 was observed in PC cells following schizandrin A treatment (Fig. 3F-G). These results suggest that schizandrin A may effectively and concurrently inhibit the activity of COX-2 and ALOX5 and induce a decrease in their expression levels.

**Figure 3.**
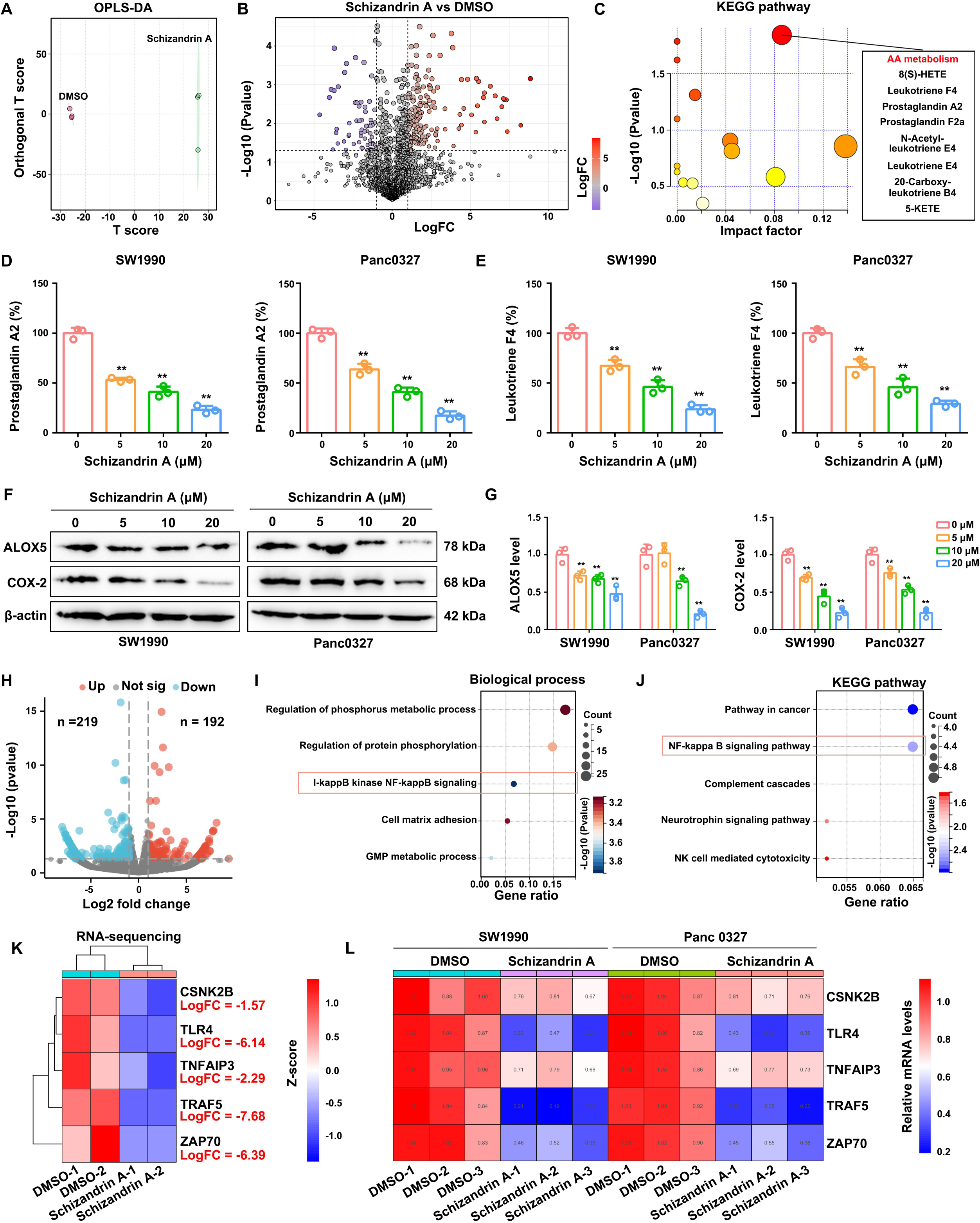
Schizandrin A inhibited the biological functions of ALOX5/COX-2 in PC cells. (A) OPLS-DA analysis of the SW1990 cells upon DMSO and Schisandrin A treatment. (B) Volcano plots demonstrate the metabolite level change in SW1990 cells after treatment with DMSO and Schisandrin A. (C) KEGG analysis for the down-regulated metabolites induced by schizandrin A. (D) ELISA experiments were performed to detect the levels of PGA2 in PC cells after treatment with schizandrin A. (E) ELISA experiments were performed to detect the levels of leukotriene F4 in PC cells after treatment with schizandrin A. (F-G) Expression of COX-2 and ALOX5 was detected in PC cells after treatment with schizandrin A. (H) Volcano plot demonstrates the differentially expressed genes in SW1990 cells after treatment with Schizandrin A. (I) GO analysis (BP terms) for down-regulated genes induced by schizandrin A. (J) KEGG analysis for down-regulated genes induced by schizandrin A. (K) Heatmap indicated the down-regulated genes induced by schizandrin A enriched in NF-κB pathway. (L) qRT-PCR was performed to detect the mRNA level change of genes enriched in NF-κB pathway after schizandrin A treatment. **, P<0.01.

Prior studies have demonstrated that prostaglandins and leukotrienes, enzymatically produced by COX-2 and ALOX5, can initiate various signaling pathways, including NF-κB, in cancer cells, thereby facilitating cell proliferation [20,21]. Consequently, we conducted an investigation into the impact of schizandrin A on the downstream pathway of COX-2 and ALOX5. Utilizing RNA-sequencing, our findings revealed that treatment of SW1990 cells with schizandrin A resulted in the up-regulation of 192 genes and the down-regulation of 219 genes (Fig. 3H). GO analysis was performed for the 219 down-regulated genes, and the results indicated that these genes were significantly enriched in “regulation of phosphorus metabolic process”, “regulation of protein phosphorylation”, “I-kappB kinase NF-kappB signaling”, “Cell matrix adhesion” and “GMP metabolic process” (Fig. 3I). Subsequently, through the implementation of KEGG enrichment analysis, it was determined that the down-regulated genes influenced by schizandrin A exhibited significant enrichment in various pathways, including “pathway in cancer”, “NF-kappaB signaling pathway”, “complement cascades”, “neurotrophin signaling pathway”, and “NK cell mediated cytotoxicity” (Fig. 3J). Specifically, five down-regulated genes, namely CSNK2B, TLR4, TNFAIP3, TRAF5, and ZAP70, were found to be enriched in the NF-κB pathway (Fig. 3K). In accordance with the findings of RNA-sequencing, qRT-PCR results demonstrated that the expression levels of CSNK2B, TLR4, TNFAIP3, TRAF5, and ZAP70 were diminished to differing extents in PC cells following treatment with schizandrin A (Fig. 3L). These observations suggested that schizandrin A may modulate the NF-κB pathway, a downstream pathway of ALOX5 and COX-2.

### 3.4. Schizandrin A inhibited the proliferation of PC cells *in vitro* and *in vivo*

Given that COX-2 and ALOX5 were identified as oncogenes in PC cells, our study sought to investigate the impact of schizandrin A on the proliferation of these cells. Utilizing the CCK-8 assay to assess cell viability in 48 h, we determined the median inhibitory concentration (IC50) of schizandrin A for HPEC, SW1990, BxPc-3, Panc0327, and CFPAC-1 cells to be 276.00 μM, 4.13 μM, 9.12 μM, 10.03 μM, and 16.12 μM, respectively (Fig. 4A). Furthermore, our findings indicated that treatment with schizandrin A (5 μM) resulted in decreased EDU incorporation rates (Fig. 4B) and reduced colony formation ability in SW1990 and Panc0327 cells (Fig. 4C).

**Figure 4.**
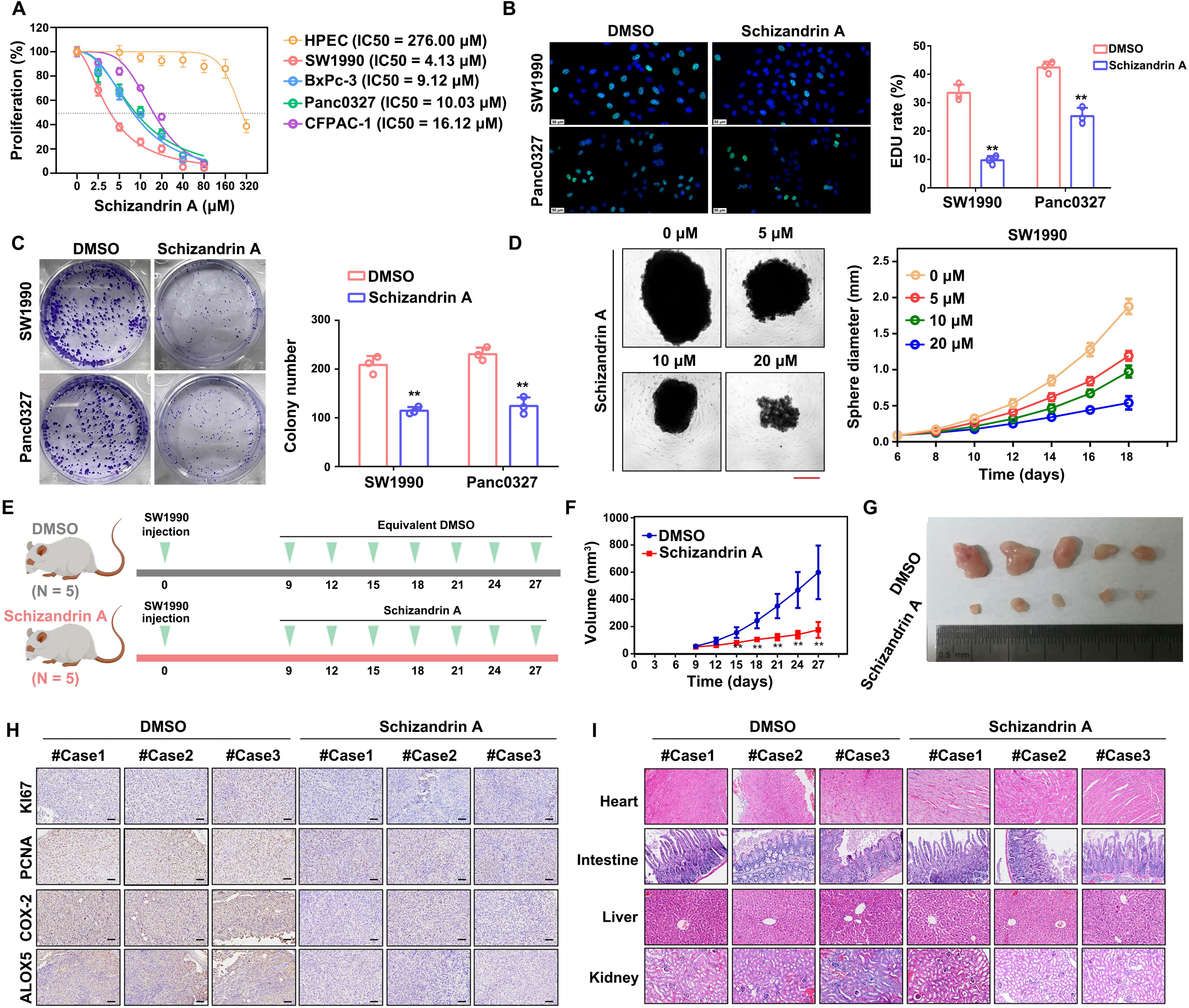
Schizandrin A inhibited the proliferation of PC cells *in vitro* and *in vivo*. (A) CCK-8 was used to detect the IC_50_ of schizandrin A for HPEC, SW1990, Panc0327, BxPc-3 and CFPAC-1 cells. (B) EDU assays were used to detect the positive rate in PC cells after schizandrin A treatment. (C) The colony formation ability was detected after schizandrin A treatment. (D) Spheroiding ability of SW1990 was detected after schizandrin A treatment. (E-G) A subcutaneous graft tumor model was utilized, revealing that tissues from the schizandrin A treatment group displayed reduced proliferation rates and tumor weight. (H) The expression of KI67, PCNA, COX-2, and ALOX5 in tumor tissues derived from cells treated with DMSO and Schizandrin A determined using IHC. (I) HE staining was performed to determine the injury of the heart, intestine, liver, and kidneys in mice treated with DMSO and Schizandrin A. **, P<0.01.

In the 3D culture model, it was observed that schizandrin A significantly impeded the spherogenesis of SW1990 cells (Fig. 4D). Additionally, a subcutaneous graft tumor model was utilized (Fig. 4E), revealing that tissues from the schizandrin A treatment group displayed reduced proliferation rates (Fig. 4F) and tumor weight (Fig. 4G). Furthermore, IHC analysis of tumor tissues indicated that the schizandrin A treatment group exhibited decreased expression of KI67, PCNA, COX-2, and ALOX5 (Fig. 4H). Furthermore, through hematoxylin and eosin (HE) staining, we found that nude mice treated with schizandrin A had no heart, liver, intestine, or kidney injury (Fig. 4I). These evidences indicated that schizandrin A exhibited prominent anti-tumor effects for PC with limited toxicity and side effects.

### 3.5. Schizandrin A suppressed the transform of normal fibroblasts (NFs) to cancer associated fibroblasts (CAFs)

Prior research has demonstrated the significance of prostaglandins and leukotrienes, which are catalyzed by COX-2 and ALOX-5, as crucial cytokines in the tumor microenvironment. These molecules play a key role in recruiting and activating various immune-related cells and stromal cells, ultimately leading to the development of an immunosuppressive microenvironment [22,23]. Given schizandrin A’s potential to inhibit the production of prostaglandins and leukotrienes, we further investigated the effects of schizandrin A on the tumor microenvironment of PC tissues. Utilizing the EPIC algorithm on PC tissues in the TCGA cohort (Fig. 5A), our analysis revealed a positive correlation between the expression of COX-2 and ALOX5 with the presence of CAFs in PC tissues (Fig. 5B). Furthermore, examination of FAP expression, a known biomarker of CAFs, in PC tissues demonstrated increased levels in tissues with high expression of COX-2 and ALOX5 (Fig. 5C-D). These findings suggest a potential association between COX-2/ALOX5 expression and the activation of CAFs in PC.

**Figure 5.**
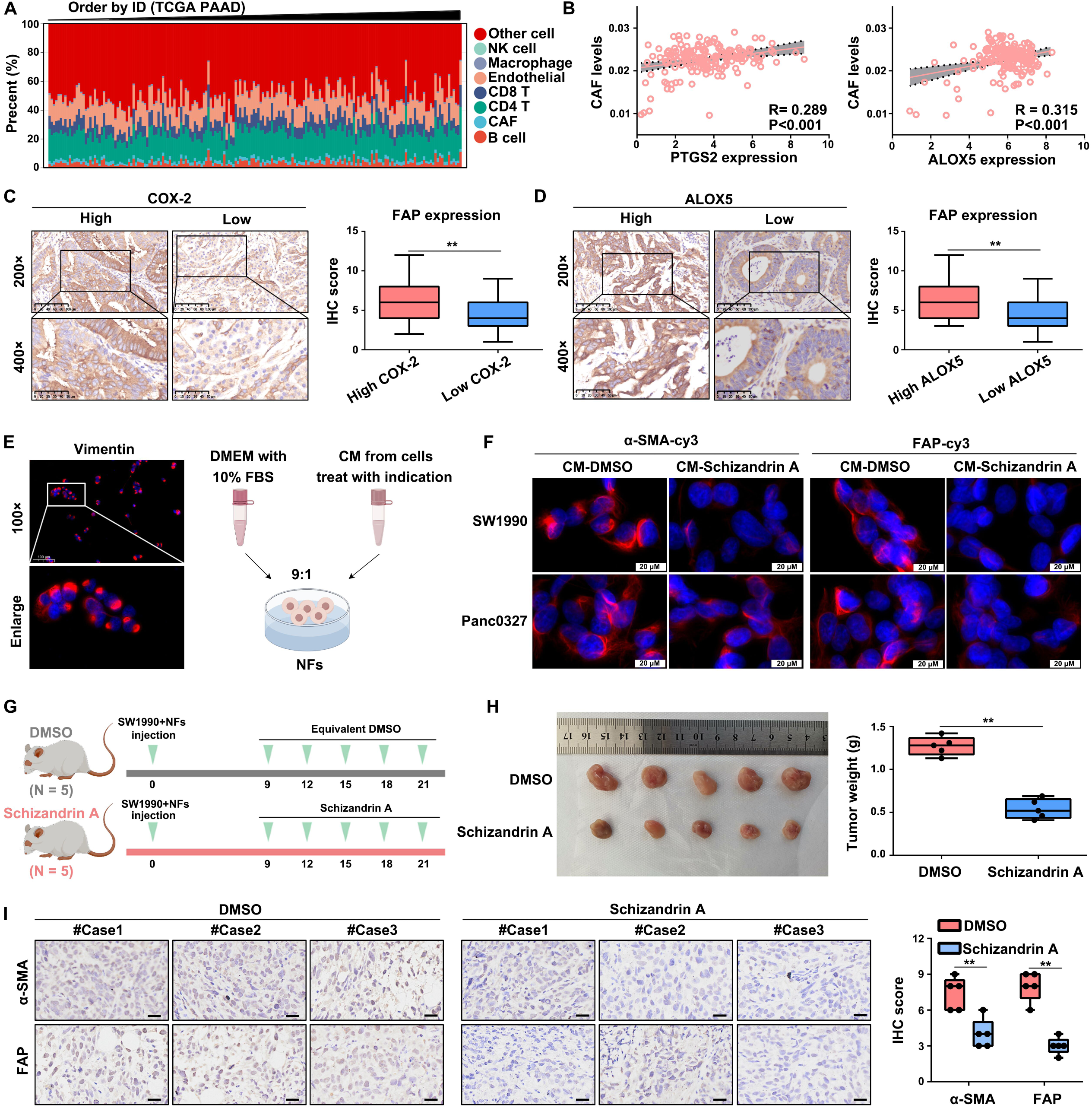
Schizandrin A suppressed the transform of normal fibroblasts (NFs) to cancer associated fibroblasts (CAFs). (A) EPIC algorithm was used to analyze immune cells in PC tissues from the TCGA cohort. (B) The positive association between COX-2/ALOX5 expression and CAF levels in PC tissues from TCGA cohort. (C) PC tissues with high COX-2 expression exhibited up-regulated FAP expression. (D) PC tissues with high ALOX5 expression exhibited up-regulated FAP expression. (E) Conditioned medium (CM) derived from PC cells treated with both DMSO or schizandrin A was collected and utilized to treat normal fibroblasts (NFs) with a 90% vimentin positive rate. (F) Immunofluorescence analysis indicated that NFs cells cultured with CM from schizandrin A treatment-PC cells exhibited lower FAP and α-SMA. (G) Mode pattern of the subcutaneous tumor formation test. (H) Tumors derived from NFs and SW1990 cells after schizandrin A demonstrated reduced tumor volume and weight. (I) Immunohistochemical analysis of tumor tissues revealed a lower expression rate of α-SMA and FAP in tumors derived from NFs and SW1990 cells treated with schizandrin A compared to those treated with DMSO. **, P<0.01.

Thus, we subsequently investigated the potential of schizandrin A to inhibit the activation of PC cells. Conditioned medium (CM) derived from PC cells treated with both DMSO or schizandrin A was collected and utilized to treat normal fibroblasts (NFs) with a 90% vimentin positive rate (Fig. 5E). The results revealed that NFs exposed to CM from PC cells treated with schizandrin A exhibited reduced levels of α-SMA and FAP expression (Fig. 5F). These findings suggest that schizandrin A has the ability to suppress the transformation of NFs into CAFs induced by PC cells *in vitro*.

Subsequently, NFs and SW1990 cells were combined and subcutaneously injected into mice (Fig. 5G). The resulting tumors derived from NFs and SW1990 cells after schizandrin A demonstrated reduced tumor volume and weight (Fig. 5H). Immunohistochemical analysis of tumor tissues revealed a lower expression rate of α-SMA and FAP in tumors derived from NFs and SW1990 cells treated with schizandrin A compared to those treated with DMSO (Fig. 5I). These findings suggested that schizandrin A has the potential to inhibit the transformation of NFs into CAFs induced by PC cells *in vivo*.

### 3.6. Mutation of the binding sites in ALXO5 (Arg246) and COX-2 (Ile124 and Ser126) inhibited the binding between schizandrin A and ALOX5/COX-2

To assess the impacts of schizandrin A on PC cells in relation to ALOX5/COX-2, mutant plasmids were generated to alter specific amino acids in ALOX5 (Arg 246) and COX-2 (Ile124 and Ser126) as Ala (Fig. 6A-B). We then transfected these wildtype (WT) and MUT plasmids into 293T cells, and we found that mutation of binding sites in COX-2 and ALOX5 did not significantly affect the catalytic activity of COX-2 and ALOX5 in 293T cells (Fig. 6C). Similarly, we found that mutation of binding sites in COX-2 and ALOX5 did not significantly affect the proliferation-promoting functions of COX-2 and ALOX5 (Fig. 6D-F).

**Figure 6.**
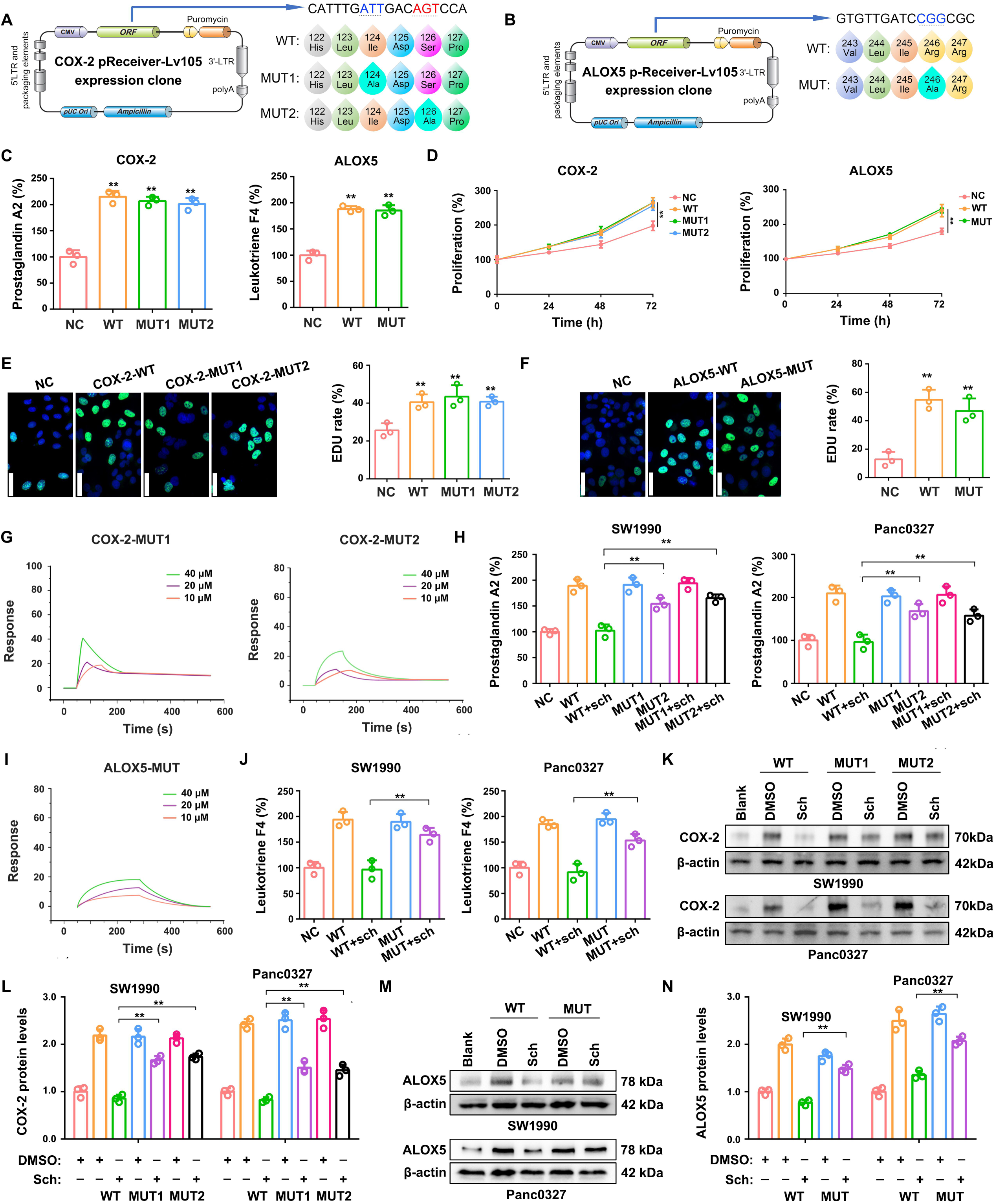
Mutation of the binding sites in ALXO5 (Arg246) and COX-2 (Ile124 and Ser126) inhibited the binding between schizandrin A and ALOX5/COX-2. (A-B) Schematic construction of COX-2- and ALOX5-expression plasmids with binding site mutations. (C) Mutation of binding sites in COX-2 and ALOX5 did not significantly affect the catalytic activity of COX-2 and ALOX5 in 293T cells. (D-F) CCK-8 and EDU assays indicated that mutation of binding sites in COX-2 and ALOX5 did not significantly affect the proliferation-promoting functions of COX-2 and ALOX5. (G) SPR experiments indicated that titration with 40 μM of schizandrin A did not reach equilibrium dissociation with the mutant COX-2 protein. (H) ELISA experiments indicated that inhibitory effects of schizandrin A on COX-2 catalytic activity were found to be substantially reversed by mutations in the binding sites. (I) SPR experiments indicated that titration with 40 μM of schizandrin A did not reach equilibrium dissociation with the mutant ALOX5 protein. (J) ELISA experiments indicated that inhibitory effects of schizandrin A on ALOX5 catalytic activity were found to be substantially reversed by mutations in the binding sites. (K-L) Schizandrin A effectively reduced the expression of WT COX-2 in PC cells, but not in cells expressing MUT proteins. (M-N) Schizandrin A effectively reduced the expression of WT ALOX5 in PC cells, but not in cells expressing MUT proteins. **, P<0.01.

However, through performing SPR, we found that, titration with 40 μM of schizandrin A did not reach equilibrium dissociation with the mutant protein, suggesting that mutations in the binding sites of COX-2 greatly diminish its affinity for schizandrin A (Fig. 6G). Furthermore, the inhibitory effects of schizandrin A on COX-2 catalytic activity were found to be substantially reversed by mutations in the binding sites, consistent with the SPR findings (Fig. 6H). Similarly, we revealed that mutation of the binding site (Arg 246) in ALOX5 resulted in a significant decrease in affinity for schizandrin A (Fig. 6I), as well as a reduction in the inhibitory effects of schizandrin A on ALOX5 catalytic activity (Fig. 6J). Additionally, transfection of WT and MUT plasmids of COX-2 and ALOX5 in SW1990 and Panc0327 cells demonstrated that schizandrin A effectively reduced the expression of WT COX-2 (Fig. 6K-L) and ALOX5 (Fig. 6M-N) in PC cells, but not in cells expressing MUT proteins. Collectively, these findings suggested that mutations in the binding sites of ALXO5 (Arg246) and COX-2 (Ile124 and Ser126) impede the interaction between schizandrin A and ALOX5/COX-2, thereby diminishing the suppressive impacts of schizandrin A on the catalytic activity and expression of ALOX5/COX-2.

### 3.7. Mutation of the binding sites in ALXO5 (Arg246) and COX-2 (Ile124 and Ser126) suppressed the inhibitory effects of schizandrin A on PC cell proliferation and CAF activation

We further determine the effects of binding site mutation on the biological functions of schizandrin A on PC and NF cells. Utilizing the CCK-8 assay, our results indicated that schizandrin A displayed a higher IC50 value in cells expressing MUT-COX2 or MUT-ALOX5 compared to those expressing WT-COX-2 or WT-ALOX5 (Fig. 7A-B). Consistent with the findings of the CCK-8 assay, the results of the EDU assay (Fig. 7C-D) and 3D sphere formation experiments (Fig. 7E-F) suggested that PC cells expressing mutant forms of COX-2 and ALOX5 protein exhibited a higher rate of EDU positivity and larger sphere volume following treatment with schizandrin A compared to cells expressing wild-type forms of these proteins. Additionally, our study demonstrated that CM from PC cells expressing mutant forms of COX-2 and ALOX5 proteins treated with schizandrin A have a greater ability to induce the transformation of NFs into CAFs, as opposed to CM from cells expressing wild-type forms of these proteins treated with schizandrin A (Fig. 7G-H). Taken together, these findings suggested that the mutation of the binding sites in ALXO5 (Arg246) and COX-2 (Ile124 and Ser126) attenuated the suppressive effects of schizandrin A on the proliferation of PC cells and activation of CAFs.

**Figure 7.**
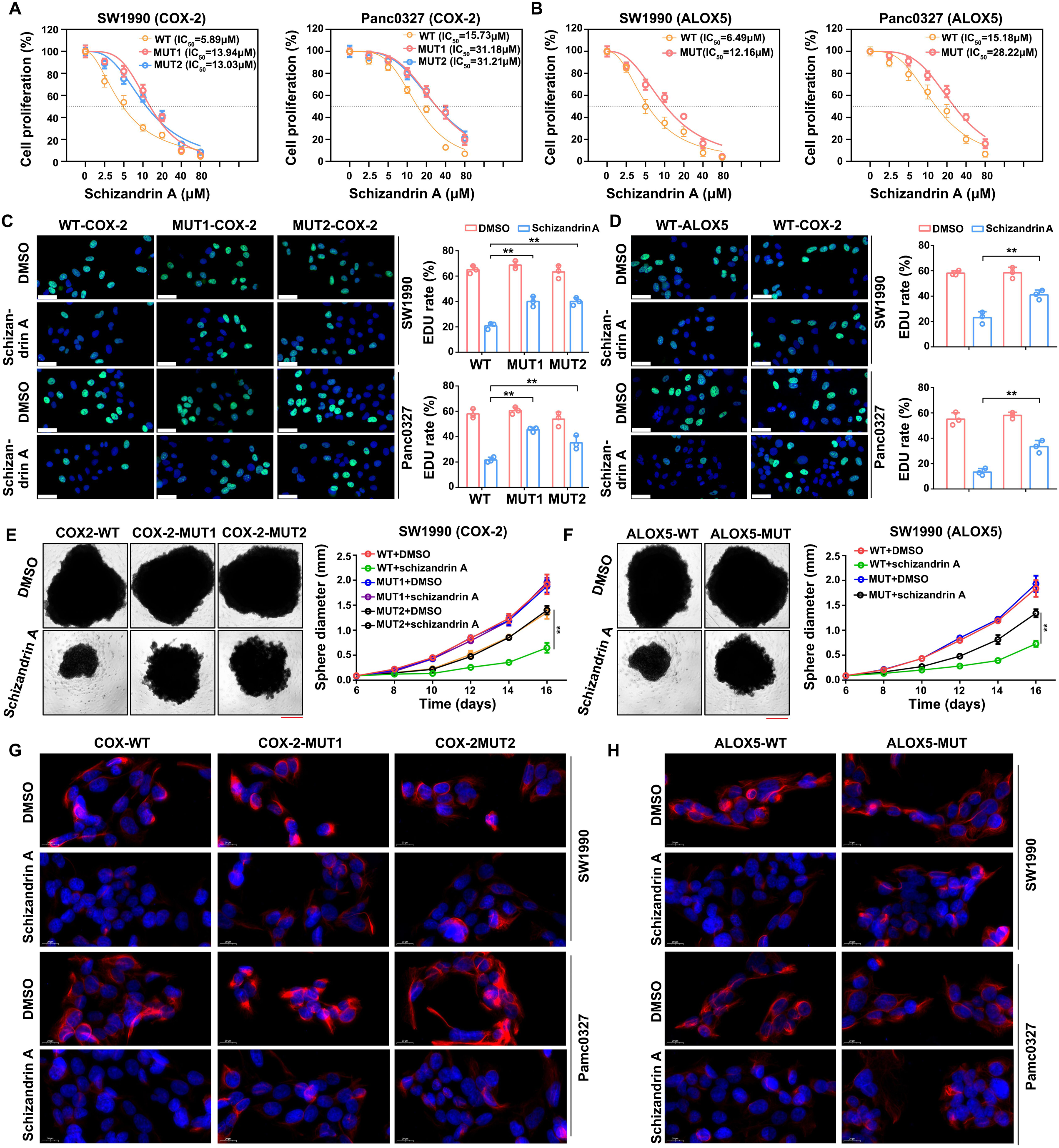
Mutation of the binding sites in ALXO5 (Arg246) and COX-2 (Ile124 and Ser126) suppressed the inhibitory effects of schizandrin A on PC cell proliferation and CAF activation. (A) PC cells expressing mutation of COX-2 protein exhibited higher IC_50_ for schizandrin A. (B) PC cells expressing mutation of ALOX5 protein exhibited higher IC_50_ for schizandrin A. (C-D) EDU assay suggested that PC cells expressing mutant forms of COX-2 and ALOX5 protein exhibited a higher rate of EDU positivity following treatment with schizandrin A compared to cells expressing wild-type forms of these proteins. (E-F) 3D sphere formation experiments suggested that PC cells expressing mutant forms of COX-2 and ALOX5 protein exhibited a larger sphere volume following treatment with schizandrin A compared to cells expressing wild-type forms of these proteins. (G-H) CM from PC cells expressing mutant forms of COX-2 and ALOX5 proteins treated with schizandrin A have a greater ability to induce the transformation of NFs into CAFs, as opposed to CM from cells expressing wild-type forms of these proteins treated with schizandrin A.**, P<0.01.

## 4. Discussion

Currently, natural products and their derivatives account for no less than 8% of all FDA-approved drugs annually. Compared with chemosynthetic drugs, the active substances obtained from natural products have novel structures and less toxic side effects [24,25]. To date, various active substances provided by natural products have been used as dual-targets in clinical tumor therapy [26,27]. For example, curcumin, a natural phenolic compound, has been used in the clinical treatment of tumors by directly targeting p300/CREB [28]. Dihydroartemisinin, a sesquiterpene lactone compound containing peroxide bridges, targets RelA/p65 and exhibits favorable anti-tumor effects in the clinical setting [29]. Schizandrin A, a ligand compound, exhibited significant hepatic protection effects and was predicted to inhibit AA metabolism as we previously reported [19]. Herein, through network pharmacology, computer molecular docking and SPR experiment, schizandrin A was found to bind to COX-2 and ALOX5 with high affinity. Therefore, schizandrin A was considered as a COX-2/ALOX5 inhibitor.

COX-2 is a key enzyme involved in AA metabolism by catalyzing AA conversion to a PG, thus activating a series of cytokine-cytokine receptor pathways, including JAK-STAT [30], TNF-α/NF-kappa B [31], and TLR [32]; therefore, high COX-2 expression generates a series of diseases, including autoimmune diseases and cancers [33,34]. Therefore, targeted COX-2 therapy has long been considered as a strategy for these diseases, and a series of COX-2 inhibitors, such as celecoxib and rofecoxib, have been developed [35,36]. However, the existence of COX-2/ALOX5 shunting has, unfortunately, limited the clinical efficacy of COX-2 inhibitors in tumor therapies [37]. Notably, not all tumors exhibit elevated COX-2/ALOX5 signatures, thus necessitating a thorough analysis of the application of schizandrin A prior to investigating its biological role.

Our current study indicated that PC tissues exhibited higher COX-2/ALOX5 signature, while high COX-2/ALOX5 signature associated poor prognosis. Furthermore, we found that no significant difference between patients with up-regulation of both ALOX5 and COX-2 compared to those with up-regulation of only ALOX5 or COX-2. However, patients with low expression of both ALOX5 and COX-2 had significantly longer survival times compared to PC patients in other groups. These results indicated that the significance of simultaneous inhibition of COX-2 and ALOX5 in the treatment of PC, and schizandrin A may be suitable for PC therapy. Consistent with the consideration, schizandrin A significantly inhibited the activity and expression of COX-2/ALOX5 in PC, as well as reducing PC cell proliferation *in vitro* and *in vivo*.

Prior research has indicated that different elements within the tumor microenvironment (TME), including hypoxia, acidosis, cytokines, and immune status, interact synergistically and contribute to the proliferation of PC cells, as well as impacting treatment outcomes [38,39]. Prostaglandins and leukotrienes, which are catalyzed by COX-2 and ALOX-5, as crucial cytokines in the tumor microenvironment [22,23]. For example, prostaglandins were showed to prevent dendritic cell accumulation and activation in tumor tissues [40]. ALOX5 was up-regulated in cholangiocarcinoma tissues, while leukotrienes catalyzed by ALOX5 promoted the recruitment of tumor associated macrophages [41]. In the current study, we found that COX-2 and ALOX5 was both associated with the levels of CAFs in PC tissues.

Fibroblasts play a crucial role in the microenvironment of solid tumors by providing structural support to the tumor tissue. As tumors progress, tumor cells release cytokines such as prostaglandins and leukotrienes, which stimulate the transformation of fibroblasts into CAFs. Subsequently, CAFs facilitate the proliferation of tumor cells through a complex network of intercellular communication [42,43]. Elevated levels of CAFs in PC tissues have been associated with a negative prognosis [44,45]. Consequently, targeting the inhibition of CAF activation may represent a promising strategy for PC therapy. Our current investigation revealed that schizandrin A effectively suppressed the transformation of normal fibroblasts into tumor-associated fibroblasts by PC cells, suggesting its potential regulatory effects within the tumor microenvironment of PC.

## Conclusion

In conclusion, we demonstrated that schizandrin A directly bound with COX-2 and ALOX5, reduced their activation and the production of leukotrienes and prostaglandins, thus exhibiting distinguished effects on suppressing PC cell proliferation and inhibiting the ability of PC cell to induce normal fibroblasts to transform into tumor-associated fibroblasts (Fig. 8).

**Figure 8.**
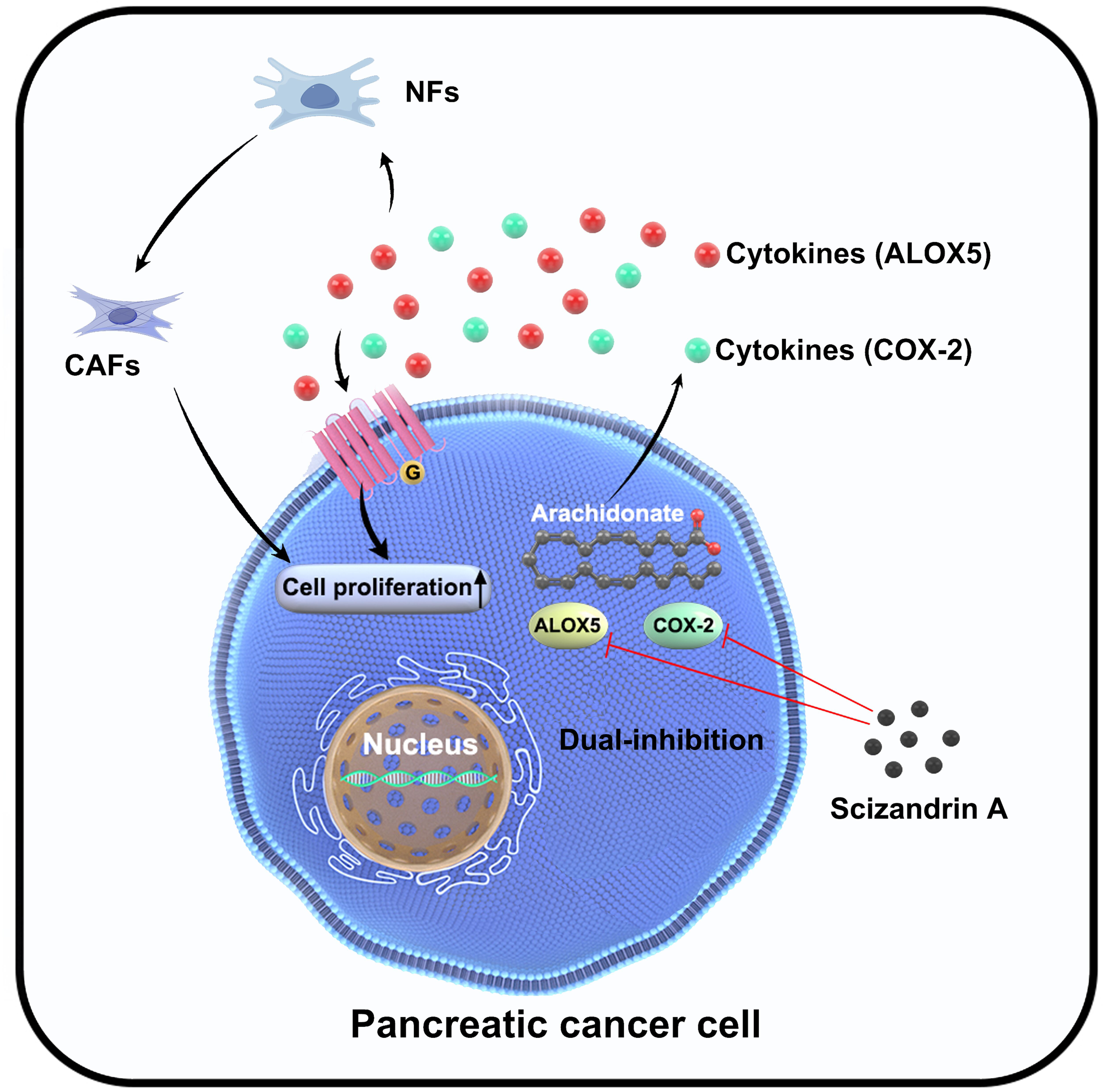
Pattern diagram demonstrated that schizandrin A directly bound with COX-2 and ALOX5, reduced their activation and the production of leukotrienes and prostaglandins, thus exhibiting distinguished effects on suppressing PC cell proliferation and inhibiting the ability of PC cell to induce normal fibroblasts to transform into tumor-associated fibroblasts.

## Declarations

## Acknowledgments

Not applicable.

## Authors’ contributions

CTX and HXJ conceived and designed the experiments. ZZR, LS, WJY and LDH performed most of the experiments and data analyses. ZZR and CTX drafted the manuscript. TQT and YYS helped with in vivo experiments. All authors read and approved the final manuscript.

## Availability of data and materials

The datasets used and/or analyzed during the current study are available from the corresponding author upon reasonable request.

## Ethics approval and consent to participate

All animal experiments were approved by the Medical Ethics Committee of Guizhou Medical University and conducted according to the guidelines for the care and use of animals for scientific research. The collection and utilization of PC specimens were approved by the Human Research Ethics Review Committee at Guizhou Medical University.

## Consent for publication

Not applicable.

## Reference

1. Kolbeinsson HM, Chandana S, Wright GP, Chung M (2023) Pancreatic Cancer: A Review of Current Treatment and Novel Therapies. J Invest Surg36:2129884.

2. Ettrich TJ, Seufferlein T (2021) Systemic Therapy for Metastatic Pancreatic Cancer. Curr Treat Options Onco22:106.

3. Wood LD, Canto MI, Jaffee EM, Simeone DM (2022) Pancreatic Cancer: Pathogenesis, Screening, Diagnosis, and Treatment. Gastroenterology163:386–402.e1.

4. Roshani R, McCarthy F, Hagemann T (2014) Inflammatory cytokines in human pancreatic cancer. Cancer Lett345:157–163.

5. Yu JH, Kim H (2014) Oxidative stress and cytokines in the pathogenesis of pancreatic cancer. J Cancer Prev19:97–102.

6. Fishbein A, Hammock BD, Serhan CN, Panigrahy D (2021) Carcinogenesis: Failure of resolution of inflammation? Pharmacol Ther 218:107670.

7. Kim W, Son B, Lee S, Do H, Youn B (2018) Targeting the enzymes involved in arachidonic acid metabolism to improve radiotherapy. Cancer Metastasis Rev37:213–225.

8. Ferrer MD, Busquets-Cortés C, Capó X, Tejada S, Tur JA, Pons A, Sureda A (2019) Cyclooxygenase-2 Inhibitors as a Therapeutic Target in Inflammatory Diseases. Curr Med Chem26:3225–3241.

9. Nagaraju GP, El-Rayes BF (2019) Cyclooxygenase-2 in gastrointestinal malignancies. Cancer 125:1221–1227.

10. Jara-Gutiérrez Á, Baladrón V(2021) The Role of Prostaglandins in Different Types of Cancer. Cells 10:1487.

11. Chen C, Guan J, Gu X, Chu Q, Zhu H (2022) Prostaglandin E2 and Receptors: Insight Into Tumorigenesis, Tumor Progression, and Treatment of Hepatocellular Carcinoma. Front Cell Dev Biol10:834859.

12. Li S, Jiang M, Wang L, Yu S (2020) Combined chemotherapy with cyclooxygenase-2 (COX-2) inhibitors in treating human cancers: Recent advancement. Biomed Pharmacother129:110389.

13. Cuzick J, Otto F, Baron JA, Brown PH, Burn J, Greenwald P, Jankowski J, La Vecchia C, Meyskens F, Senn HJ, Thun M (2009) Aspirin and non-steroidal anti-inflammatory drugs for cancer prevention: an international consensus statement. Lancet Oncol 10:501–507.

14. Ganesh R, Marks DJ, Sales K, Winslet MC, Seifalian AM (2012) Cyclooxygenase/lipoxygenase shunting lowers the anti-cancer effect of cyclooxygenase-2 inhibition in colorectal cancer cells. World J Surg Oncol 10:200.

15. Park SW, Heo DS, Sung MW (2012) The shunting of arachidonic acid metabolism to 5-lipoxygenase and cytochrome p450 epoxygenase antagonizes the anti-cancer effect of cyclooxygenase-2 inhibition in head and neck cancer cells. Cell Oncol (Dordr) 35:1–8.

16. Atanasov AG, Zotchev SB, Dirsch VM (2021) International Natural Product Sciences Taskforce; Supuran CT. Natural products in drug discovery: advances and opportunities. Nat Rev Drug Discov20:200–216.

17. Sowndhararajan K, Deepa P, Kim M, Park SJ, Kim S (2018) An overview of neuroprotective and cognitive enhancement properties of lignans from Schisandra chinensis. Biomed Pharmacother97:958–968.

18. Rybnikář M, Šmejkal K, Žemlička M (2019) Schisandra chinensis and its phytotherapeutical applications. Ceska Slov Farm68:95–118.

19. Hong M, Zhang Y, Li S, Tan HY, Wang N, Mu S, Hao X, Feng Y(2017)A Network Pharmacology-Based Study on the Hepatoprotective Effect of Fructus Schisandrae. Molecules 22:1617.

20. Khan MA, Khan MJ (2018) Nano-gold displayed anti-inflammatory property via NF-kB pathways by suppressing COX-2 activity. Artif Cells Nanomed Biotechnol 46:1149–1158.

21. Cheng JH, Zhang WJ, Zhu JF, Cui D, Song KD, Qiang P, Mei CZ, Nie ZC, Ding BS, Han Z, Ding ZE, Zheng WW (2021) CaMKIIγ regulates the viability and self-renewal of acute myeloid leukaemia stem-like cells by the Alox5/NF-κB pathway. Int J Lab Hematol43:699–706.

22. Wang D, Cabalag CS, Clemons NJ, DuBois RN (2021) Cyclooxygenases and Prostaglandins in Tumor Immunology and Microenvironment of Gastrointestinal Cancer. Gastroenterology161:1813–1829.

23. Tian W, Jiang X, Kim D, Guan T, Nicolls MR, Rockson SG (2020) Leukotrienes in Tumor-Associated Inflammation. Front Pharmacol11:1289.

24. Newman DJ, Cragg GM (2016) Natural Products as Sources of New Drugs from 1981 to 2014. J Nat Prod79: 629–661.

25. Dong S, Guo X, Han F, He Z, Wang Y (2022) Emerging role of natural products in cancer immunotherapy. Acta Pharm Sin B12:1163–1185.

26. Wang Y, Yang L, Hou J, Zou Q, Gao Q, Yao W, Yao Q, Zhang J (2019) Hierarchical virtual screening of the dual MMP-2/HDAC-6 inhibitors from natural products based on pharmacophore models and molecular docking. J Biomol Struct Dyn37:649–670.

27. Hernández ÁP, Díez P, García PA, Pérez-Andrés M, Ortega P, Jambrina PG, Díez D, Castro MÁ, Fuentes M(2020)A Novel Cytotoxic Conjugate Derived from the Natural Product Podophyllotoxin as a Direct-Target Protein Dual Inhibitor. Molecules25:4258.

28. Balasubramanyam K, Varier RA, Altaf M, Swaminathan V, Siddappa NB, Ranga U, Kundu TK(2004)Curcumin, a novel p300/CREB-binding protein-specific inhibitor of acetyltransferase, represses the acetylation of histone/nonhistone proteins and histone acetyltransferase-dependent chromatin transcription. J Biol Chem279:51163–51171.

29. Li N, Sun W, Zhou X, Gong H, Chen Y, Chen D, Xiang F(2019) Dihydroartemisinin Protects against Dextran Sulfate Sodium-Induced Colitis in Mice through Inhibiting the PI3K/AKT and NF-κB Signaling Pathways. Biomed Res Int2019:1415809.

30. Rai A, Kumar U, Raj V, Singh AK, Kumar P, Keshari AK, Kumar D, Maity B, De A, Samanta A, et al(2018)Novel 1,4-benzothazines obliterate COX-2 mediated JAK-2/STAT-3 signals with potential regulation of oxidative and metabolic stress during colorectal cancer. Pharmacol Res132:188–203.

31. Plummer R, Hu GF, Liu T, Yoo J(2020)Angiogenin regulates PKD activation and COX-2 expression induced by TNF-α and bradykinin in the colonic myofibroblast. Biochem Biophys Res Commun 525:870–876.

32. Echizen K, Hirose O, Maeda Y, Oshima M(2016)Inflammation in gastric cancer: Interplay of the COX-2/prostaglandin E2 and Toll-like receptor/MyD88 pathways. Cancer Sci107:391–397.

33. Chen W, Zhong Y, Feng N, Guo Z, Wang S, Xing D (2021) New horizons in the roles and associations of COX-2 and novel natural inhibitors in cardiovascular diseases. Mol Med27:123.

34. Li S, Jiang M, Wang L, Yu S(2020) Combined chemotherapy with cyclooxygenase-2 (COX-2) inhibitors in treating human cancers: Recent advancement. Biomed Pharmacother 129:110389.

35. Tołoczko-Iwaniuk N, Dziemia ń czyk-Pakieła D, Nowaszewska BK, Celi ń ska-Janowicz K, Miltyk W(2019)Celecoxib in Cancer Therapy and Prevention - Review. Curr Drug Targets 20:302–315.

36. Ziesenitz VC, Welzel T, van Dyk M, Saur P, Gorenflo M, van den Anker JN (2022) Efficacy and Safety of NSAIDs in Infants: A Comprehensive Review of the Literature of the Past 20 Years. Paediatr Drugs24:603–655.

37. Gautam S, Roy S, Ansari MN, Saeedan AS, Saraf SA, Kaithwas G (2017) DuCLOX-2/5 inhibition: a promising target for cancer chemoprevention. Breast Cancer24:180–190.

38. Chen D, Zhang X, Li Z, Zhu B (2021) Metabolic regulatory crosstalk between tumor microenvironment and tumor-associated macrophages. Theranostics11:1016–1030.

39. de Visser KE, Joyce JA(2023)The evolving tumor microenvironment: From cancer initiation to metastatic outgrowth. Cancer Cell 41:374–403.

40. Zelenay S, van der Veen AG, Böttcher JP, Snelgrove KJ, Rogers N, Acton SE, Chakravarty P, Girotti MR, Marais R, Quezada SA, et al(2015) Cyclooxygenase-Dependent Tumor Growth through Evasion of Immunity. Cell162:1257–1270.

41. Chen J, Tang Y, Qin D, Yu X, Tong H, Tang C, Tang Z(2023)ALOX5 acts as a key role in regulating the immune microenvironment in intrahepatic cholangiocarcinoma, recruiting tumor-associated macrophages through PI3K pathway. J Transl Med21:923.

42. Lavie D, Ben-Shmuel A, Erez N, Scherz-Shouval R(2022)Cancer-associated fibroblasts in the single-cell era. Nat Cance3:793–807.

43. Chhabra Y, Weeraratna AT(2023)Fibroblasts in cancer: Unity in heterogeneity. Cell. 186:1580–1609.

44. Qi R, Bai Y, Li K, Liu N, Xu Y, Dal E, Wang Y, Lin R, Wang H, Liu Z, et al (2023) Cancer-associated fibroblasts suppress ferroptosis and induce gemcitabine resistance in pancreatic cancer cells by secreting exosome-derived ACSL4-targeting miRNAs. Drug Resist Updat 68:100960.

45. Pan X, Zhou J, Xiao Q, Fujiwara K, Zhang M, Mo G, Gong W, Zheng L (2021) Cancer-associated fibroblast heterogeneity is associated with organ-specific metastasis in pancreatic ductal adenocarcinoma. J Hematol Oncol14:184.

